# Preventive and Need-Driven Use of Dental Services among Community-Dwelling Older People with Long-Term Care Needs

**DOI:** 10.64898/2026.01.29.26345104

**Authors:** Tomoko Wakui, Ayako Edahiro, Tsuyoshi Okamura, Yoshiko Motohashi, Takumi Hirata, Hirohiko Hirano

## Abstract

Oral health is a key component of quality of life among older adults, yet maintaining preventive dental care becomes increasingly difficult for those with long-term care (LTC) needs. This study examined patterns of preventive and need-driven dental service use among home-dwelling older adults with LTC needs in Japan and identified the caregiving, functional, and socioeconomic factors associated with these patterns. Data were drawn from a nationwide online survey of family caregivers reporting dental service use among older adults certified under the Japanese Long-Term Care Insurance system (n = 1,055).

Poorer financial situations increased the likelihood of a lack of dental visits, whereas need-driven visits were more common among those who received intensive or long-term caregiving and whose caregivers reported a greater burden. Dementia status was not independently associated with dental service use. These findings highlight the importance of promoting preventive dental care within community-based LTC systems, particularly as care needs intensify.

**Key Points:**

- Financial strain was linked to having no dental visits
- More intensive and longer caregiving, especially with higher caregiver burden, was linked to need-driven dental visits
- Dementia status was not independently associated with dental service use

## Introduction

Maintaining oral health is often framed as an issue of primary health care or dental medicine. However, among people living with long-term care (LTC) needs, oral health is deeply embedded in daily life, caregiving practices, and access to community support systems, rather than being solely a clinical concern (Tokyo Metropolitan Institute for Geriatrics and Gerontology, 2021). As care needs increase, individuals gradually experience more difficulty in performing oral self-care, recognizing problems in the oral cavity, and initiating appropriate care-seeking behaviors (Gluzman et al., 2013; Ornstein et al., 2015). Consequently, oral health may deteriorate, interfering with enjoyable eating experiences and contributing to undernutrition and frailty (Hussein et al., 2022; Tanaka et al., 2017). These processes highlight the importance of a preventive approach to oral health care that emphasizes regular monitoring and early intervention rather than reliance on symptom-driven dental visits only after problems become apparent (Gaewkhiew et al., 2017; Medina-Solis et al., 2019; Mohd Khairuddin et al., 2024; Montero et al., 2014; Thomson et al., 2010).

In Japan, where 29.7% of the population is aged 65 years and older and the number of people living with LTC needs continues to increase, the Community-based Integrated Care System (chiiki hōkatsu care system) (Ministery of Health Labour and Welfare; Tsutsui, 2014) has long been promoted to enable older adults to continue living in familiar communities through the coordinated provision of medical care, LTC, preventive services, housing, and daily life support. As the number of people living with dementia has increased beyond what existing care frameworks alone could accommodate, Japan introduced the Basic Act on Dementia to Promote an Inclusive Society in 2023 (“Basic Act on Dementia to Promote the Realization of a Symbiotic Society”, 2023; Ministry of Health Labour and Welfare, 2023) as a system-level response to this demographic and social shift. Building on the foundations of community-based integrated care, the Comprehensive Strategy for the Promotion of Dementia Policies (Ministry of Health Labour and Welfare) emphasizes the timely and appropriate provision of medical and LTC services according to the stage of care needs, with the overarching goal of enabling people living with LTC needs—particularly those living with dementia—to live with dignity and continuity in familiar community settings.

Within this broader policy landscape, oral health has been explicitly positioned as a component of dementia-responsive care. National guidelines recommend continuous oral health management aligned with the progression of cognitive decline, and efforts have been made to strengthen the role of dental professionals through dementia-specific training programs for dentists (Tokyo Metropolitan Institute for Geriatrics and Gerontology, 2021). These initiatives highlight expectations for the dental sector not only in the early detection of oral and cognitive changes but also in the ongoing management of oral health among older adults with LTC needs living at home. Taken together, these policy initiatives indicate a shift toward preventive and continuous oral health management across the dementia trajectory rather than episodic, problem-driven dental care.

Despite these policy-level efforts, there is little empirical evidence available concerning how dental services are actually utilized in everyday life by home-dwelling older adults with LTC needs, including those living with dementia. In particular, little is known about the real-world utilization of dental services among home-dwelling older adults who require LTC, including those living with dementia. Existing policy discussions tend to emphasize system readiness and professional capacity, but the extent to which older adults and their families are able to access preventive dental care in community settings has not been sufficiently examined.

Oral health is a critical component of overall health and well-being, particularly among older adults and individuals living with dementia (Tokyo Metropolitan Institute for Geriatrics and Gerontology, 2021). Maintaining good oral hygiene can significantly influence quality of life (Zini & Sgan-Cohen, 2008), enabling better nutrition (Hussein et al., 2022), communication, and social interactions (Cicciù et al., 2013; Scambler et al., 2023). However, there are numerous challenges in ensuring optimal oral health within this population, including cognitive decline, behavioral issues, practical difficulties in providing oral care, and complex decision-making regarding dental treatment (Curtis et al., 2021; Geddis-Regan et al., 2024; Idris et al., 2024; Scambler et al., 2023).

Research has indicated that people with dementia often experience poorer oral health outcomes relative to the general elderly population, which is attributed to issues such as difficulties in managing daily oral hygiene routines, limited access to dental services, and inadequate training of caregivers (Curtis et al., 2021; Kc et al., 2021; Scambler et al., 2023; Soilemezi et al., 2023; Yamaguchi et al., 2021). Caregivers and healthcare professionals frequently report challenges in assessing oral health needs and providing effective interventions, which are compounded by fragmented healthcare services and a lack of integrated oral health promotion strategies (Kc et al., 2021; Soilemezi et al., 2023). Emanuel reported that despite a high rate of dental attendance among patients with early-stage dementia, preventive care—such as oral hygiene education and fluoride supplementation—was insufficiently provided (Emanuel & Sorensen, 2018). Specifically, only approximately 20% of patients reported receiving dietary or oral hygiene advice, and only 14% had received fluoride supplementation, indicating an underutilization of preventive strategies that are essential for maintaining oral health in this vulnerable group.

Furthermore, exploring the perspectives of carers and health professionals reveals the need for improved communication, continuous training, and policy changes to prioritize oral health within dementia care frameworks. Studies highlight the importance of oral health education and support for family caregivers to improve their confidence and skills in managing oral health, thereby positively impacting the dental health of older people (Manchery et al., 2020; Soilemezi et al., 2023).

Previous studies have highlighted substantial challenges in accessing dental care among older adults with cognitive decline. However, much of the existing evidence is derived from qualitative research (Curtis et al., 2021; Scambler et al., 2023; Soilemezi et al., 2023), which has primarily documented perceived barriers to obtaining dental services and the experiences of older adults and their families when seeking care. While these studies provide important insights into factors that may hinder dental access, the extent to which routine dental check-ups are actually utilized in real-world community settings remains insufficiently examined. Moreover, improving oral health among people living with dementia requires attention to the complex interplay of individual functional limitations, family caregiving arrangements, and service accessibility within LTC systems. Addressing these challenges calls for coordinated approaches that integrate dental services into broader community-based care, involving both formal and informal caregivers. To inform such efforts, empirical evidence on patterns of dental service use is needed to support effective service planning and policy development.

However, dental service use among home-dwelling older adults with LTC needs—including those living with dementia—remains understudied. In particular, limited evidence exists regarding how regularly people with dementia are able to access dental services and which care-related factors are associated with their use of dental services in community settings. Clarifying real-world patterns of dental service use, including both preventive and problem-driven use, and related factors may inform equitable and accessible community oral health planning. From a preventive care perspective, regular dental check-ups represent an opportunity for early detection and ongoing management of oral health problems(Emanuel & Sorensen, 2018; Kc et al., 2021), whereas need-driven dental visits typically occur after symptoms interfere with daily life. This study aimed to investigate patterns of dental service use—including preventive and need-driven visits—among home-dwelling older adults with LTC needs and to examine how these patterns vary according to their caregiving arrangements, functional care needs, socioeconomic conditions, and dementia status.

## Method

### Study design

This study employed a cross-sectional design to examine dental service use behavior among community-dwelling older adults with LTC needs in Japan. A nationwide online survey was conducted with family caregivers, who provided proxy information on the dental service use of older adults living at home and who were certified as requiring care under the Japanese Long-Term Care Insurance (LTCI) system. Data were collected from 1,166 family caregivers registered with an online survey panel managed by MyVoice Communications, Inc.(MyVoice Communications Inc.).

The eligibility criteria for the study participants were as follows: (1) being aged 20 years or older; (2) self-reporting the provision of care to at least one community-dwelling older adult aged 65 years or older who was certified as requiring care under Japan’s Long-Term Care Insurance (LTCI) system; and (3) not being institutionalized themselves. The present questionnaire survey was conducted as part of a follow-up wave of an online caregiver cohort established in 2023, and all participants had been providing care since at least 2023.

### Instruments

The primary outcome of this study was the dental service use behavior of community-dwelling older adults with LTC needs, which was assessed through caregiver reports. Annual dental service use was classified to reflect established distinctions in dental epidemiology between routine or regular dental check-ups (preventive-oriented), problem-oriented or symptom-driven visits, and no dental visits during the past year (Camargo et al., 2012; Gaewkhiew et al., 2017; Thomson et al., 2010; Yoshino et al., 2016). In this framework, regular check-ups is considered the optimal preventive approach, whereas no visits represents a potentially problematic pattern, as it reflects missed opportunities for prevention, early detection, and timely management of oral health conditions.

The care recipient characteristics included age, dementia status, relationship with the caregiver (spouse/partner, parent, parent-in-law, or other), and coresidence with the caregiver. Functional status was assessed using six activities of daily living (ADLs: bathing, dressing, toileting, mobility, continence, and eating) and six instrumental activities of daily living (IADLs: meal preparation, shopping, housekeeping and laundry, mobility outside the home, medication management, and money management). Each item was rated on a three-point scale (0 = fully dependent, 1 = partially dependent, 2 = independent), with higher scores indicating greater functional independence (Katz et al., 1963; Lawton & Brody, 1969).

The caregiving context variables included the frequency of caregiving per week and duration of caregiving. Caregiver characteristics included sex, age, marital status, employment status, perceived financial situation, self-rated health, and caregiving burden. Caregiving burden was measured using the Japanese version of the Zarit Caregiver Burden Inventory (ZBI) (Arai et al., 1997).

In addition, dental care–related factors included whether the care recipient had a regular personal dentist and the type of dental service use (clinic-based care, home-visit dental care, or both).

### Data analysis

Descriptive analyses were first conducted to summarize dental service use behavior and participant characteristics. Analyses were restricted to respondents with complete data on the outcome and all relevant explanatory variables (n = 1,055). Bivariate analyses were subsequently performed to examine the associations between dental service use behavior and care recipient characteristics, caregiver characteristics, caregiving context, functional status, and dental care–related factors. Chi-square tests were applied to categorical variables, and analysis of variance (ANOVA) was used for continuous variables, as appropriate.

To identify factors independently associated with dental service use patterns, multinomial logistic regression analyses were conducted using regular dental check-ups as the reference category. All variables related to care recipient characteristics, caregiver characteristics, caregiving context, functional status, and dental care system factors were considered for inclusion in the multivariable models. All statistical analyses were performed using IBM SPSS Statistics for Windows, version 29.0, and *p* < .05 was considered to indicate statistical significance.

### Ethical considerations

This study was ethically approved by the Tokyo Metropolitan Institute for Geriatrics and Gerontology Institutional Review Board (No. R21-076). This study complied with the Declaration of Helsinki and its amendments or comparable ethical standards for conducting the survey. Informed consent was obtained from all participants via online methods.

## Results

Table 1 summarizes the sociodemographic characteristics and caregiving situations by dental service use behavior among older adults with LTC needs. The analyzed sample included 1,055 community-dwelling older adults with LTC needs, with a mean age of 84.7 years (SD = 7.7). Approximately two-thirds of care recipients had dementia and lived with their caregivers.

**Table 1.** Sociodemographic Characteristics and Caregiving Situations by Dental Checkup Behavior (N = 1055)

| Variables | No visits, n<br>(%)<br>n = 317 | Regular<br>checkups, n (%)<br>n = 367 | Need-driven<br>visits, n (%)<br>n = 371 | Total | P value |
| --- | --- | --- | --- | --- | --- |
| Relationship |  |  |  |  | 0.002<sup>a</sup> |
| Mother | 19 | 17.3 | 19.5 | 55.8 |  |
| Father | 4.3 | 6.5 | 8 | 18.8 |  |
| Mother-in-law | 2.9 | 3.1 | 3.1 | 9.2 |  |
| Father-in-law | 0.4 | 1.3 | 0.7 | 2.4 |  |
| Other | 2.2 | 3.1 | 1.9 | 7.2 |  |
| Spouse/Partner | 1.3 | 3.3 | 2 | 6.6 |  |
| CR age | 85.7 ± 7.43 | 83.6 ± 8.05 | 85.1 ± 7.43 | 84.7 ± 7.69 | n.s.<sup>b</sup> |
| Dementia |  |  |  |  | n.s.<sup>a</sup> |
| Yes | 21 | 22 | 22.3 | 65.3 |  |
| None | 9 | 12.8 | 12.9 | 34.7 |  |
| ADL (1 = Fully<br>dependent-3 =<br>Independent) |  |  |  |  |  |
| Bathing | 1.63 ± 0.77 | 1.67 ± 0.72 | 1.84 ± 0.78 | 1.72 ± 0.76 | < 0.001<sup>b</sup> |
| Dressing | 1.93 ± 0.76 | 1.97 ± 0.71 | 2.13 ± 0.72 | 2.01 ± 0.74 | < 0.001<sup>b</sup> |
| Toileting | 2.03 ± 0.83 | 2.08 ± 0.80 | 2.27 ± 0.79 | 2.13 ± 0.81 | < 0.001<sup>b</sup> |
| Mobility | 1.69 ± 0.68 | 1.8 ± 0.69 | 1.88 ± 0.69 | 1.79 ± 0.69 | 0.003<sup>b</sup> |
| Continence | 2.02 ± 0.83 | 2.15 ± 0.79 | 2.28 ± 0.76 | 2.16 ± 0.80 | < 0.001<sup>b</sup> |
| Eating | 1.32 ± 0.59 | 1.45 ± 0.65 | 1.52 ± 0.72 | 1.44 ± 0.67 | < 0.001<sup>b</sup> |
| IADL (1: Fully<br>dependent-3:<br>Independent) |  |  |  |  |  |
| Meal preparation | 2.36 ± 0.78 | 2.36 ± 0.75 | 2.52 ± 0.68 | 2.42 ± 0.74 | 0.004<sup>b</sup> |
| Shopping | 1.14 ± 0.40 | 1.32 ± 0.55 | 1.31 ± 0.58 | 1.26 ± 0.53 | < 0.001<sup>b</sup> |
| Housekeeping &<br>laundry | 1.22 ± 0.52 | 1.39 ± 0.63 | 1.43 ± 0.70 | 1.36 ± 0.63 | < 0.001<sup>b</sup> |
| Mobility outside | 1.19 ± 0.45 | 1.31 ± 0.51 | 1.35 ± 0.56 | 1.29 ± 0.52 | < 0.001<sup>b</sup> |
| Medication<br>management | 1.41 ± 0.68 | 1.72 ± 0.79 | 1.68 ± 0.76 | 1.61 ± 0.76 | < 0.001<sup>b</sup> |
| Money management | 1.34 ± 0.64 | 1.56 ± 0.75 | 1.58 ± 0.75 | 1.50 ± 0.72 | < 0.001<sup>b</sup> |
| Coresidence |  |  |  |  | 0.061<sup>a</sup> |
| Live-in | 20.5 | 20.8 | 22.9 | 64.2 |  |
| Live-out | 9.6 | 14 | 12.2 | 35.8 |  |
| Care frequency |  |  |  |  | 0.043<sup>a</sup> |
| Everyday | 17.7 | 16.2 | 16.8 | 50.7 |  |
| 5-6/week | 1.7 | 3.5 | 2.8 | 8.1 |  |
| 2-4/week | 6.1 | 8.1 | 8.3 | 22.5 |  |
| 1 day/week | 2.6 | 3.3 | 3.2 | 9.1 |  |
| Less than 1day/week | 2 | 3.7 | 4 | 9.7 |  |
| Caregiving period |  |  |  |  | 0.009<sup>a</sup> |
| ≤ 1 year | 2.8 | 4.5 | 2.5 | 9.8 |  |
| 1–3 years | 7.9 | 11.6 | 10.6 | 30 |  |
| 3–5 years | 7.2 | 8.3 | 10.9 | 26.4 |  |
| 5–10 years | 8.2 | 7 | 8 | 23.2 |  |
| ≥ 10 years | 3.9 | 3.4 | 3.2 | 10.5 |  |
| CG sex |  |  |  |  | n.s.<sup>a</sup> |
| Female | 13.5 | 14.8 | 17 | 45.2 |  |
| Male | 16.6 | 20 | 18.2 | 54.8 |  |
| CG age | 57.0 ± 9.55 | 56.1 ± 10.88 | 57.2 ± 9.47 | 56.8 ± 10.01 | 0.001<sup>b</sup> |
| Marital status |  |  |  |  | < 0.001<sup>a</sup> |
| Unmarried | 11.5 | 8.4 | 10.9 | 30.8 |  |
| Married | 15.6 | 24.2 | 21.3 | 61.1 |  |
| Divorced/Deceased | 2.9 | 2.2 | 2.9 | 8.1 |  |
| Employment status |  |  |  |  |  |
| Part-time/Other | 5 | 4.5 | 6.4 | 15.9 |  |
| Full-time | 13.7 | 21.1 | 18.2 | 53.1 |  |
| Unemployed | 11.3 | 9.1 | 10.6 | 31 |  |
| Financial situation |  |  |  |  | 0.011<sup>a</sup> |
| Very comfortable | 1.9 | 2.8 | 2.2 | 6.9 |  |
| Somewhat comfortable | 6.9 | 11.6 | 9.8 | 28.2 |  |
| Neither | 8.4 | 8.9 | 9.2 | 26.5 |  |
| Somewhat strained | 6.6 | 6.9 | 9.4 | 22.9 |  |
| Very strained | 6.2 | 4.5 | 4.6 | 15.4 |  |
| Self-rated health |  |  |  |  | n.s.<sup>a</sup> |
| Very healthy | 13.5 | 14.8 | 17 | 45.2 |  |
| Fairly healthy | 16.6 | 20 | 18.2 | 54.8 |  |
| Neither healthy nor unhealthy | 5.3 | 5.2 | 6.4 | 17 |  |
| Not very healthy | 6 | 6.9 | 7.4 | 20.3 |  |
| Not healthy at all | 2.2 | 1.1 | 2 | 5.3 |  |
| Caregiving burden | 12.8 ± 8.89 | 11.1 ± 7.81 | 12.6 ± 8.77 | 12.1 ± 8.51 | 0.011<sup>b</sup> |
| Regular personal dentist |  |  |  |  | 0.043<sup>a</sup> |
| Yes | – | 49.2 | 42.6 | 91.8 |  |
| No | – | 0.8 | 7.4 | 8.2 |  |
| Type of dental service use |  |  |  |  | < 0.001<sup>a</sup> |
| Clinic-based | – | 33.7 | 40.4 | 74.1 |  |
| Home-visit | – | 14.9 | 8.8 | 23.7 |  |
| Both | – | 1.1 | 1.1 | 2.2 |  |

*Note.* Values are presented as % or mean ± SD. Percentages are calculated using the total sample size as the denominator, except for the ‘Total’ column, which presents column percentages.
<sup>a</sup> Tested using the chi-square test.
<sup>b</sup> Tested using analysis of variance (ANOVA).

The mean age of the caregivers was 56.8 years (SD = 10.0), they were slightly more often male, and most were married and employed. Financial situations and caregiving burden varied across the sample. Most care recipients had a regular personal dentist, and clinic-based dental care was the most common service type, followed by home-visit dental services.

No significant differences were observed in dementia status across groups. In contrast, ADL and IADL functioning showed consistent and significant gradients: greater levels of independence in all ADL domains (bathing, dressing, toileting, mobility, continence, and eating) and IADL domains (meal preparation, shopping, housekeeping, mobility outside, medication management, and money management) were associated with regular or need-driven dental visits rather than no visits (*p* < .01).

Among caregiver characteristics, caregiver age (*p* = .001), marital status (*p* < .001) and employment status (*p* = .001) were also associated with dental service use behavior: regular check-ups were more common among younger, married and full-time employed caregivers. Financially strained households (*p* = .011) were more likely to have no dental visits. Caregivers with higher burden scores were observed in the no-visit and need-driven visit groups compared with those reporting regular use of dental services (*p* = .011). Dental service use behavior differed significantly across caregiver–care recipient relationships (*p* = .002).

The results of the multinomial logistic regression analysis examining factors associated with oral checkup behavior among older adults with LTC needs are presented in Table 2, with regular use of dental services as the reference category. For no dental visits, independence in medication management (OR = 0.53, *p* < .01) and full-time caregiver employment (OR = 0.47, *p* < .01) were associated with lower odds, reflecting a greater likelihood of maintaining regular dental service use among older adults with higher self-management capacity in medication. A worse financial situation was associated with higher odds of no visits (OR = 1.20, *p* = .02), reflecting financial barriers to dental access.

**Table 2.**
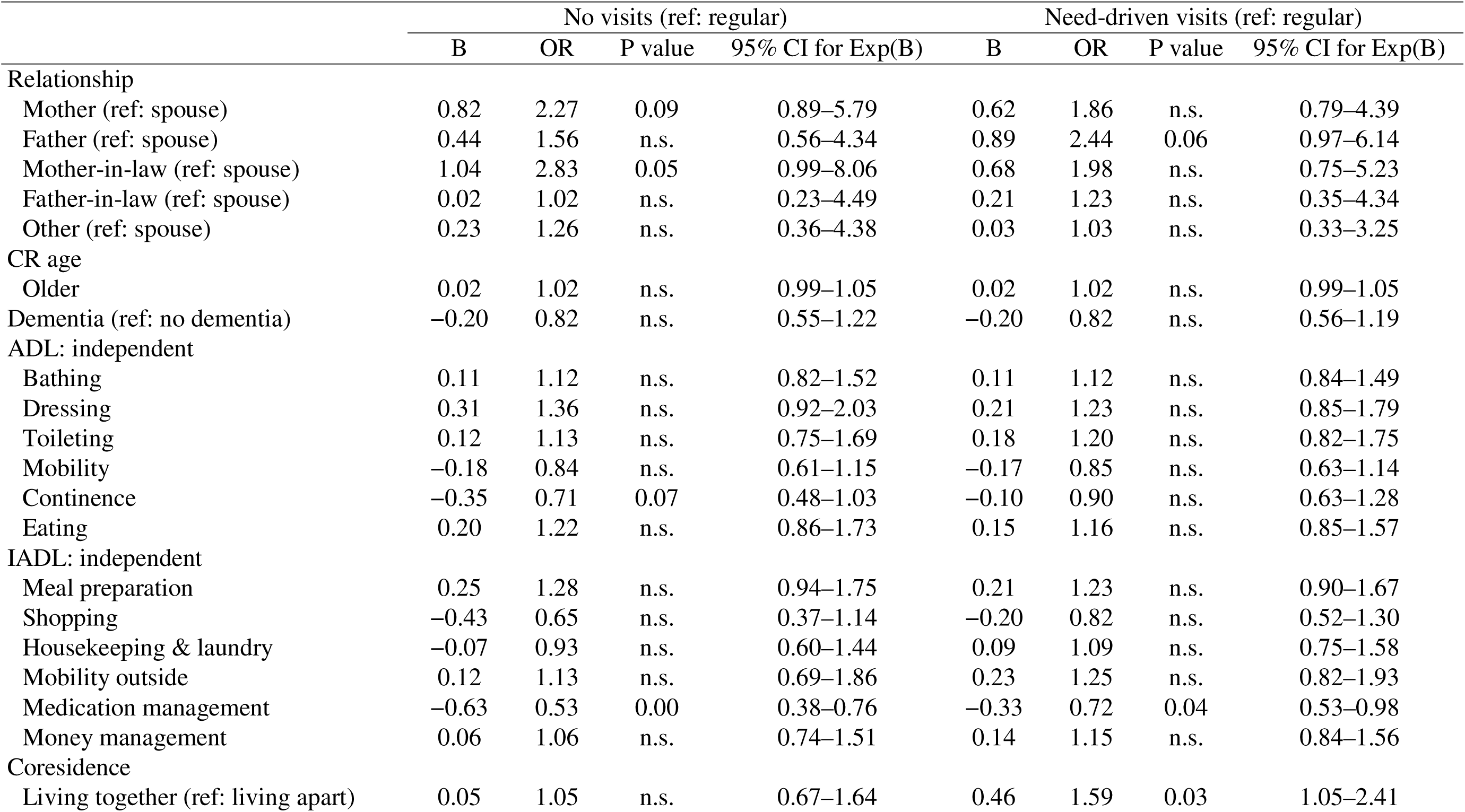
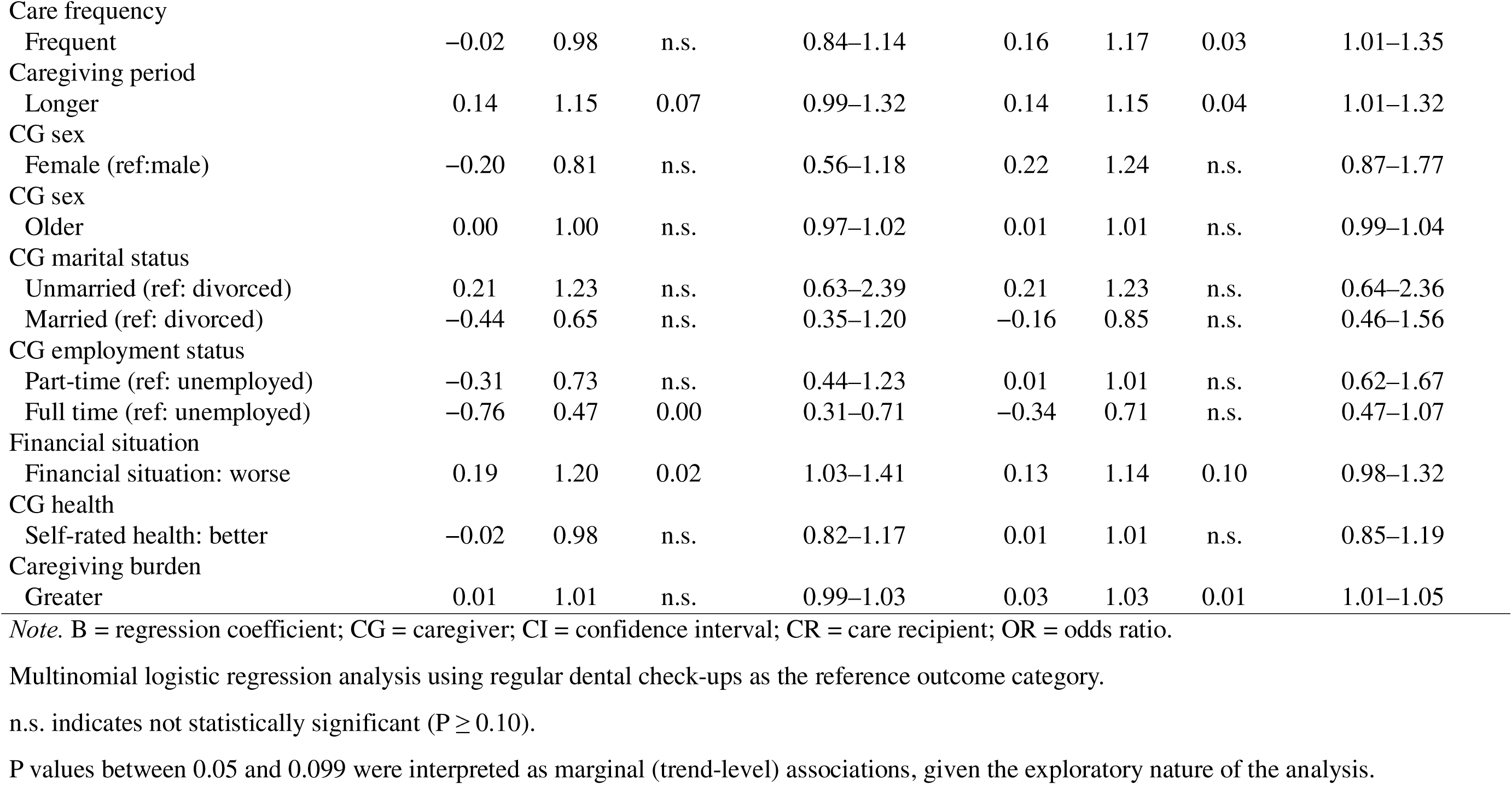
Multinomial Logistic Regression Analysis on Oral Check Up among Older Adults with Long-term Care Needs.

|  | No visits (ref: regular) |  |  |  | Need-driven visits (ref: regular) |  |  |  |
| --- | --- | --- | --- | --- | --- | --- | --- | --- |
|  | B | OR | P value | 95% CI for Exp(B) | B | OR | P value | 95% CI for Exp(B) |
| Relationship |  |  |  |  |  |  |  |  |
| Mother (ref: spouse) | 0.82 | 2.27 | 0.09 | 0.89–5.79 | 0.62 | 1.86 | n.s. | 0.79–4.39 |
| Father (ref: spouse) | 0.44 | 1.56 | n.s. | 0.56–4.34 | 0.89 | 2.44 | 0.06 | 0.97–6.14 |
| Mother-in-law (ref: spouse) | 1.04 | 2.83 | 0.05 | 0.99–8.06 | 0.68 | 1.98 | n.s. | 0.75–5.23 |
| Father-in-law (ref: spouse) | 0.02 | 1.02 | n.s. | 0.23–4.49 | 0.21 | 1.23 | n.s. | 0.35–4.34 |
| Other (ref: spouse) | 0.23 | 1.26 | n.s. | 0.36–4.38 | 0.03 | 1.03 | n.s. | 0.33–3.25 |
| CR age |  |  |  |  |  |  |  |  |
| Older | 0.02 | 1.02 | n.s. | 0.99–1.05 | 0.02 | 1.02 | n.s. | 0.99–1.05 |
| Dementia (ref: no dementia) | −0.20 | 0.82 | n.s. | 0.55–1.22 | −0.20 | 0.82 | n.s. | 0.56–1.19 |
| ADL: independent |  |  |  |  |  |  |  |  |
| Bathing | 0.11 | 1.12 | n.s. | 0.82–1.52 | 0.11 | 1.12 | n.s. | 0.84–1.49 |
| Dressing | 0.31 | 1.36 | n.s. | 0.92–2.03 | 0.21 | 1.23 | n.s. | 0.85–1.79 |
| Toileting | 0.12 | 1.13 | n.s. | 0.75–1.69 | 0.18 | 1.20 | n.s. | 0.82–1.75 |
| Mobility | −0.18 | 0.84 | n.s. | 0.61–1.15 | −0.17 | 0.85 | n.s. | 0.63–1.14 |
| Continence | −0.35 | 0.71 | 0.07 | 0.48–1.03 | −0.10 | 0.90 | n.s. | 0.63–1.28 |
| Eating | 0.20 | 1.22 | n.s. | 0.86–1.73 | 0.15 | 1.16 | n.s. | 0.85–1.57 |
| IADL: independent |  |  |  |  |  |  |  |  |
| Meal preparation | 0.25 | 1.28 | n.s. | 0.94–1.75 | 0.21 | 1.23 | n.s. | 0.90–1.67 |
| Shopping | −0.43 | 0.65 | n.s. | 0.37–1.14 | −0.20 | 0.82 | n.s. | 0.52–1.30 |
| Housekeeping & laundry | −0.07 | 0.93 | n.s. | 0.60–1.44 | 0.09 | 1.09 | n.s. | 0.75–1.58 |
| Mobility outside | 0.12 | 1.13 | n.s. | 0.69–1.86 | 0.23 | 1.25 | n.s. | 0.82–1.93 |
| Medication management | −0.63 | 0.53 | 0.00 | 0.38–0.76 | −0.33 | 0.72 | 0.04 | 0.53–0.98 |
| Money management | 0.06 | 1.06 | n.s. | 0.74–1.51 | 0.14 | 1.15 | n.s. | 0.84–1.56 |
| Coresidence |  |  |  |  |  |  |  |  |
| Living together (ref: living apart) | 0.05 | 1.05 | n.s. | 0.67–1.64 | 0.46 | 1.59 | 0.03 | 1.05–2.41 |
| Care frequency |  |  |  |  |  |  |  |  |
| Frequent | −0.02 | 0.98 | n.s. | 0.84–1.14 | 0.16 | 1.17 | 0.03 | 1.01–1.35 |
| Caregiving period |  |  |  |  |  |  |  |  |
| Longer | 0.14 | 1.15 | 0.07 | 0.99–1.32 | 0.14 | 1.15 | 0.04 | 1.01–1.32 |
| CG sex |  |  |  |  |  |  |  |  |
| Female (ref: male) | −0.20 | 0.81 | n.s. | 0.56–1.18 | 0.22 | 1.24 | n.s. | 0.87–1.77 |
| CG sex |  |  |  |  |  |  |  |  |
| Older | 0.00 | 1.00 | n.s. | 0.97–1.02 | 0.01 | 1.01 | n.s. | 0.99–1.04 |
| CG marital status |  |  |  |  |  |  |  |  |
| Unmarried (ref: divorced) | 0.21 | 1.23 | n.s. | 0.63–2.39 | 0.21 | 1.23 | n.s. | 0.64–2.36 |
| Married (ref: divorced) | −0.44 | 0.65 | n.s. | 0.35–1.20 | −0.16 | 0.85 | n.s. | 0.46–1.56 |
| CG employment status |  |  |  |  |  |  |  |  |
| Part-time (ref: unemployed) | −0.31 | 0.73 | n.s. | 0.44–1.23 | 0.01 | 1.01 | n.s. | 0.62–1.67 |
| Full time (ref: unemployed) | −0.76 | 0.47 | 0.00 | 0.31–0.71 | −0.34 | 0.71 | n.s. | 0.47–1.07 |
| Financial situation |  |  |  |  |  |  |  |  |
| Financial situation: worse | 0.19 | 1.20 | 0.02 | 1.03–1.41 | 0.13 | 1.14 | 0.10 | 0.98–1.32 |
| CG health |  |  |  |  |  |  |  |  |
| Self-rated health: better | −0.02 | 0.98 | n.s. | 0.82–1.17 | 0.01 | 1.01 | n.s. | 0.85–1.19 |
| Caregiving burden |  |  |  |  |  |  |  |  |
| Greater | 0.01 | 1.01 | n.s. | 0.99–1.03 | 0.03 | 1.03 | 0.01 | 1.01–1.05 |

*Note.* B = regression coefficient; CG = caregiver; CI = confidence interval; CR = care recipient; OR = odds ratio.
Multinomial logistic regression analysis using regular dental check-ups as the reference outcome category.
n.s. indicates not statistically significant ( $P \geq 0.10$ ).
P values between 0.05 and 0.099 were interpreted as marginal (trend-level) associations, given the exploratory nature of the analysis.

For need-driven dental visits, living together with the care recipient (OR = 1.59, *p* = .03), more frequent caregiving (OR = 1.17, *p* = .03), a longer caregiving period (OR = 1.15, *p* = .04), and a greater caregiving burden (OR = 1.03, *p* = .01) were associated with higher odds, reflecting a shift toward symptom-driven utilization under closer and more intensive caregiving circumstances. Independence in medication management was associated with lower odds of need-driven visits (OR = 0.72, *p* = .04), reflecting a greater tendency toward regular dental service use among older adults with higher functional independence in medication management.

## Discussion

This study examined patterns of dental service utilization—both preventive and need-driven—among home-dwelling older adults with long-term care (LTC) needs in Japan and investigated caregiving, functional, and socioeconomic factors associated with utilization, including dementia status. By incorporating comprehensive information on older adults’ daily living situations, such as caregiving arrangements, this study provides a nuanced understanding of dental service use behavior in community-based LTC settings.

In contrast to previous studies that have emphasized difficulties in accessing dental care among people living with dementia (Curtis et al., 2021; Kc et al., 2021; Scambler et al., 2023; Soilemezi et al., 2023), our findings showed that dementia status itself was not associated with dental service use patterns. One possible explanation is the context of Japan’s universal medical and LTC insurance systems. In Japan, both access to healthcare services and basic literacy regarding the importance of regular dental check-ups may be relatively widespread, even among individuals living with dementia. Moreover, dental health services are integrated into the universal health insurance system, allowing access to dental care at lower costs in Japan than in other OECD countries (Aida et al., 2021). This is attributable to Japan’s long-term and sustained commitment to dental health policy (Sato et al., 2023). Moreover, dental care for older adults—including those requiring LTC and those living with dementia—has been actively promoted through national health policies in Japan (Japanese Society of Gerodontology, 2015; Miura, 2019). As a result, dementia may not function as a direct barrier to dental service utilization in the same way reported in other countries or care systems.

In addition, the study population was limited to older adults certified as needing care under the Japanese LTC insurance program, thereby focusing on individuals with ongoing support needs who are already embedded within public service systems. This inclusion criterion likely selected individuals and families who have some familiarity with healthcare and social service systems—not limited to dental care—potentially reducing disparities in access (Curtis et al., 2021) related to cognitive status.

Another possible explanation relates to the perspective of the respondents. This study relied on family caregivers as proxies for older adults requiring LTC, meaning that all participants had at least some degree of family involvement. National statistics indicate that approximately 16% of older adults receiving care report having no informal caregiver as their primary support provider (Ministry of Health Labour and Welfare, 2022). Such situations—where older adults lack informal caregiving support—were not captured in this study and may represent contexts in which dementia-related barriers to dental care are more pronounced. In particular, for older adults without family involvement or those not yet well integrated into formal care systems, dental care may be given lower priority than other immediate daily living or medical support needs (Curtis et al., 2021; Idris et al., 2024; Schluter et al., 2021; Soilemezi et al., 2023), resulting in delayed or symptom-driven dental service use.

Our multinomial logistic regression analysis revealed that, after controlling for other factors, dependence on medication management was associated with different dental service use patterns, highlighting medication management as a potentially distinctive indicator compared with other ADL/IADL measures. Dependency in medication management was associated with higher odds of having no dental visits or relying on symptom-driven dental care than regular dental check-ups. This association likely reflects a broader capacity to maintain stable daily routines and engage in preventive health behaviors. Independence in medication management reflects not only functional ability but also the presence of organizational skills, health awareness, and routine adherence that facilitate scheduling, attending, and prioritizing regular dental service use (Wang et al., 2025). In contrast, when medication management depends on caregivers, oral health care may become secondary to more immediate caregiving demands, leading dental visits to occur primarily in response to symptoms rather than as part of routine preventive care (Curtis et al., 2021; Emanuel & Sorensen, 2018).

First, these findings highlight the potential role of prescription drug services as points of early detection and referral for oral health care. Because community pharmacies interact regularly with older adults—often prior to overt functional decline—they may serve as accessible entry points for identifying early risks and providing guidance on preventive dental services. Previous studies have emphasized that closer interprofessional collaboration between pharmacists and dental professionals may encourage pharmacists to take a more active role in promoting oral self-care and facilitating referrals for both routine and problem-oriented dental care (Lygre et al., 2017). Consistent with this perspective, recent practice-based initiatives in Japan have shown that pharmacists conducting basic oral function checks in community and home-care settings were able to identify oral health problems and successfully link individuals to dental services, supporting the feasibility of pharmacy–dental collaboration for early detection and prevention (Yakuyomi Editorial Team, 2024).

More critically, primary care physicians, particularly home-visiting physicians, may play a critical role in integrating oral health considerations into ongoing medical care (Lewis et al., 2015). Home-visiting physicians frequently care for older adults with chronic conditions such as diabetes, dementia, Parkinson’s disease, or cerebrovascular disease, in whom compromised oral hygiene is a clinically relevant medical issue, as these conditions are commonly associated with dysphagia, periodontal disease, and an increased risk of pulmonary or aspiration pneumonia (Huang et al., 2025; Khadka et al., 2021; Manger et al., 2017; Umemoto & Furuya, 2020). Given their continuous and holistic involvement in patients’ daily health management, visiting physicians may be well positioned to recognize oral health risks and initiate referrals to dental services. Strengthening collaboration between visiting medical care and dental services may therefore enhance preventive care and contribute to more comprehensive coordination within the community-based integrated care system.

Financial situation also emerged as a significant barrier to dental service use among older adults with LTC needs. This finding is consistent with previous research showing that financial constraints inhibit access to medical care (Alaazi et al., 2022; Anderson & Kim, 2009; Idris et al., 2024; Wang et al., 2024). Notavly, although dental treatment in Japan is available at a coparatively lower cost than in many other OECD countries (Aida et al., 2021), financial situation remained significantly associated with dental service utilization. This indicates that barriers may reflect not only service fees but also indirect costs, such as transportation and commuting expenses, opportunity loss due to taking time off work, and the cost of arranging an accompanying person for dental visits. Accordingly, financial support mechanisms (e.g., subsidies or publicly supported programs) may be needed to reduce barriers to preventive dental care among older adults with LTC needs.

Finally, dental service use in home care settings was shaped more strongly by caregiver involvement and care arrangements than by clinical characteristics alone, consistent with the findings of previous studies (Ferguson, 2024; Manchery et al., 2020; Zenthöfer et al., 2016). Greater caregiver coresidence, more frequent caregiving, and longer caregiving period—indicators of greater caregiving involvement—were associated with need-driven rather than preventive dental service use (Emanuel & Sorensen, 2018). This pattern suggests that in intensive caregiving contexts, oral care is often addressed reactively in response to oral symptoms that affect oral intake, such as pain or functional problems, rather than proactively through regular preventive dental visits. Such findings align with the realities of dementia caregiving, where immediate daily care demands may take precedence over preventive oral health management (Curtis et al., 2021; Emanuel & Sorensen, 2018). This pattern may indicate that daily oral care is relatively well maintained by family caregivers on the basis of perceived needs. However, this approach is inherently limited. As cognitive decline progresses, older adults with dementia become increasingly unable to recognize, articulate, or report oral health problems. In this context, reliance on symptom-driven dental care is insufficient and may result in delayed detection of preventable oral conditions. These findings strongly underscore the necessity of regular, preventive dental care for people living with dementia (Edahiro et al., 2021), not merely as a response to overt symptoms but as a critical mechanism for early identification, risk management, and timely intervention before oral problems adversely affect nutrition, comfort, and overall health.

## Limitations

Although this was a nationwide study focusing on people living with dementia and LTC needs, the data were collected through family caregivers. As a result, dental check-up situations among individuals without available family caregivers may have been overlooked. To better capture the experiences of those living alone or without family support, future research could incorporate data collection through home-visiting dentists and other community-based outreach pathways that directly reach home-dwelling individuals with dementia and care needs without family members.

Furthermore, detailed oral health conditions were not assessed. For example, information on the presence of dental caries and the use of dentures or complete dentures was not collected. Such clinical indicators could provide important insights for tailoring dental check-up needs among people living with dementia (Ornstein et al., 2015; Schluter et al., 2021). Patterns of dental service use within the household may influence whether routine dental care is initiated or maintained, suggesting that shared health behaviors could affect dental service utilization among older adults with care needs.

In addition, financial circumstances warrant further consideration. While poorer financial situations were associated with reduced dental service use in this study, individuals receiving public assistance in Japan are exempt from out-of-pocket medical and dental costs. This suggests that financial barriers to dental service use may operate through mechanisms beyond direct service fees, such as transportation costs, competing daily expenses, or challenges in navigating care systems. Future studies should therefore examine how different forms of financial vulnerability—including but not limited to income level and public assistance status—interact with access to dental services among home-dwelling older adults with LTC needs.

## Acknowledgments

The authors would like to express their sincere appreciation to the family caregivers who participated in this study and generously shared their experiences. The authors are also grateful to Professor Ichiro Kai (The University of Tokyo, Graduate School of Medicine) for his insightful guidance on the policy implications of our findings, especially in relation to improving dental service access and community-based support for older adults with long-term care needs.

## Funding details

This work was supported by the Health and Welfare Services for the Elderly Program (Grant Number R6-No. 72) funded by the Ministry of Health, Labour and Welfare of Japan, a Grant-in-Aid for Scientific Research JP23K21580 and JP22K10306 from the Japan Society for the Promotion of Science.

## Declaration of interests statement

The authors report that there are no competing interests to declare.

## Data availability statement

Data are available on request because of privacy/ethical restrictions.

## Use of artificial intelligence

An AI-based tool was used to assist with English language editing and improve clarity. The AI tool was not used to generate research content, analyze data, or draw conclusions. The authors take full responsibility for the content of this manuscript.

## Notes

### Competing Interest Statement

The authors have declared no competing interest.

